# Psychosis and Associated Factors among Adolescents and Young Adults at LUTH in Zambia

**DOI:** 10.1101/2025.06.04.25329009

**Authors:** Chileleko Siakabanze, David N. Masta, Lukundo Siame, Chakulya Martin, Hanzooma Hatwiko, Emmanuel Luwaya, Joreen P. Povia, Sepiso K. Masenga

## Abstract

**Background:** Psychosis poses a significant burden in sub-Saharan Africa, yet data on risk factors in Zambia remains scarce. Understanding the factors associated with psychosis among adolescents and young adults is critical for effective intervention strategies.

**Methods:** We conducted a cross-sectional study to determine the correlates of psychosis at Livingstone university teaching hospital (LUTH). We collected sociodemographic and clinical variables. Psychosis was the outcome variable, while independent variables included age, sex, residence, employment, marital status, developmental milestones, depression, anxiety, substance use (cannabis, alcohol, and opioids), criminal history, and hematologic and liver biochemical markers. We conducted both descriptive and inferential analyses using statcrunch.

**Results:** The median age (with interquartile range) was comparable between participants diagnosed with psychosis (21 (19–23) and those without psychosis (20 (17–23). Of the study population (n=427), 84% (n=199) of participants with psychosis were male and 16% (n=38) were female. On multivariable analysis, the following variables were significantly associated with psychosis; Poor insight into mental illness was strongly associated with reduced odds of psychosis (AOR: 0.34, 95% CI 0.18, 0.64, p=0.0007), and opioid use was inversely associated (AOR: 0.43, 95%CI 0.19, 0.98, p=0.045).

**Conclusion:** This study highlights Zambia’s high burden of psychosis among young males and underscores cannabis as a modifiable risk factor. The paradoxical protective role of opioids warrants further investigation. Our findings emphasize the need for context-specific interventions, such as mental health literacy programs and harm-reduction strategies targeting substance use. Addressing systemic gaps in Zambia’s mental health infrastructure and integrating culturally sensitive diagnostic tools are critical to mitigating psychosis-related outcomes.

## Background

Psychosis, a debilitating mental health condition characterized by a disconnection from reality, poses significant challenges to individuals, families, and healthcare systems globally [1,2]. It is a multifactorial disorder influenced by genetic, environmental, and socioeconomic determinants [2]. Adolescents and young adults are particularly vulnerable, as the onset of psychotic disorders such as schizophrenia frequently occurs during this critical developmental period [3,4]. Globally, psychosis affects approximately 3% of the population, with profound implications for educational attainment, employment, and social functioning [4,5]. Meanwhile the sub-Saharan Africa (SSA) prevalence of psychosis ranges from 3.9% to 8% [6].

Multiple risk factors have been implicated in the development of psychosis; urban residence, substance use (such as cannabis and opioids), and low educational attainment among others [7,8]. Despite advances in understanding its etiology and treatment in high-income countries, there’s dearth of data from SSA to guide policy and clinical interventions [9]. Existing studies in SSA highlight unique challenges, such as the role of traditional healers in care pathways and the impact of infectious diseases like HIV on mental health, which are rarely addressed in global frameworks [10,11]. Zambia, like many SSA nations, faces a silent crisis in mental healthcare, with psychosis often undiagnosed, stigmatized, or inadequately managed [12,13]. In addition, Zambia has fewer psychiatrists serving its population of approximately 19 million, and mental health receives less than 1% of the national health budget [14]. Adolescents and young adults, who constitute over 60% of Zambia’s population, are especially at risk due to high rates of substance use, unemployment, and exposure to trauma [15].

The scarcity of robust epidemiological data on psychosis in Zambia hinders effective public health responses [16]. At LUTH, clinicians anecdotally report rising cases of psychosis among youths, yet the prevalence, demographic patterns, and associated risk factors remain unquantified. This knowledge gap has dire consequences: without evidence, policymakers cannot allocate resources effectively, and healthcare providers lack guidance for contextually relevant interventions [17]. Furthermore, cultural beliefs attributing psychosis to spiritual causes often delay biomedical treatment, leading to advanced illness stages at presentation [18]. Compounding this issue, Zambia’s mental health infrastructure is fragmented, with LUTH serving as one of the few referral centers for complex cases [13]. The hospital’s role as a teaching institution also underscores the need for localized research to train future healthcare workers in evidence-based practices [19]. Without understanding the sociodemographic and clinical correlates of psychosis in this setting, efforts to reduce its burden will remain fragmented and ineffective [20].

Furthermore, the study is justified by two interrelated factors (developmental outcomes of the affected persons, and rising substance abuse rates) [21]. Severity of developmental consequences of untreated psychotic young people, such as disrupted education, social exclusion, and long-term disability, perpetuating cycles of poverty [3,21]. In Zambia, social safety nets are weak, these outcomes place immense strain on families and communities [12]. Even though substance use is a modifiable risk factor among young adults and adolescents, there’s emerging evidence of substance use; 49% of cannabis use, 24.3% of alcohol abuse [22], 60% of inhalants and solvents such as glue sniffing [12,23] and emerging substances such codeine-containing cough syrups, heroin, and methamphetamine use are rising, particularly in urban centers like Lusaka and Livingstone [15,24].

Therefore, this study, conducted at LUTH, aimed at addressing established gaps by examining the factors associated with psychosis among adolescents and young adults in Zambia, a population disproportionately affected by systemic inequities.

## Methodology

### Study Design

This cross-sectional study utilized retrospective programmatic data extracted from medical records at LUTH. The analysis was based on secondary data from the project *“Prevalence and Factors Associated with Cannabis Use Among Adolescents and Young Adults at LUTH in Zambia,”* which is currently undergoing peer review (Manuscript ID: PONE-D-25-23402).

### Study Setting

The study was conducted at LUTH, a tertiary referral hospital serving Zambia’s Southern Province. The psychiatry department’s records were utilized due to their systematic documentation of psychosis and substance use disorders in adolescent and young adult populations.

### Eligibility and Recruitment

We screened medical records of 1,500 adolescents and young adults (aged 10–27 years) who received care between 2022 and 2024. Of these, 427 files met the inclusion criteria and were analysed, while 1,073 were excluded due to incomplete data such as missing age, sex, residential information, or diagnosis.

### Data Collection

We conducted a review of medical records between February 1, 2025, and March 15, 2025, utilizing trained research assistants to extract the necessary data.

### Study variables

The primary outcome variable in this study was psychosis. Psychosis is a state where an individual experiences a significant disconnection from reality, often involving distortions in thinking, perception, and behaviour. The term is not a diagnosis itself, but rather a broad description of symptoms that can be found in various psychiatric and medical conditions. Diagnostic criteria (DSM-IV to DSM-5 Psychotic Disorders) vary depending on the specific disorder, but generally involve the presence of hallucinations, delusions, disorganized thinking or speech, or grossly disorganized or catatonic behaviour [25–28].

The independent variables were categorized into two domains; sociodemographic and clinical. Sociodemographic variables included age, sex, place of residence (urban or rural), employment status, and history of criminal activity. Clinical variables encompassed developmental milestone status, level of insight into mental illness, substance use (alcohol, cannabis and opioids), and psychiatric comorbidities such as depression, and anxiety. Additional clinical parameters included serum creatinine levels, alanine aminotransferase (ALT), white blood cell (WBC) count, neutrophil count, lymphocyte count, monocyte count, red blood cell (RBC) count, haemoglobin concentration, and platelet count.

### Data Analysis

The data was exported from the REDCap platform to Microsoft Excel 2013 for cleaning and coding. Subsequent analysis was conducted using Statcrunch. Categorical variables were summarized using frequencies and percentages, and continuous variables were described using medians and interquartile ranges (IQR). The Shapiro-Wilk test was utilized to assess normality of the data distribution. Associations between categorical variables were examined via the chi-square test, and differences in medians were analysed using the Wilcoxon rank-sum test. To determine factors linked to psychosis, both univariable and multivariable logistic regression models were applied.

### Ethics approval and consent to participants

The study obtained ethical clearance from the Mulungushi University School of Medicine and Health Sciences Research Ethics Committee (ethics approval number SMHS-MU2-2024-67) on June 6, 2024, and approval to access patient records was granted by the LUTH administration. All collected and analysed data were de-identified to maintain confidentiality, with no personally identifiable information recorded during data abstraction or analysis. The project utilized secondary data, and as such, written or verbal consent was deemed unnecessary and formally waived by the ethics committee.

The Strengthening the Reporting of Observational Studies in Epidemiology (STROBE) guidelines were followed to ensure methodological rigor.

## Results

From 2500 available files for abstraction, we reviewed 1500 file. A total of 427 were analysed, while 1073 were excluded due to incomplete and or missing data such as age, sex, residential information and diagnosis.

**Figure 1.**
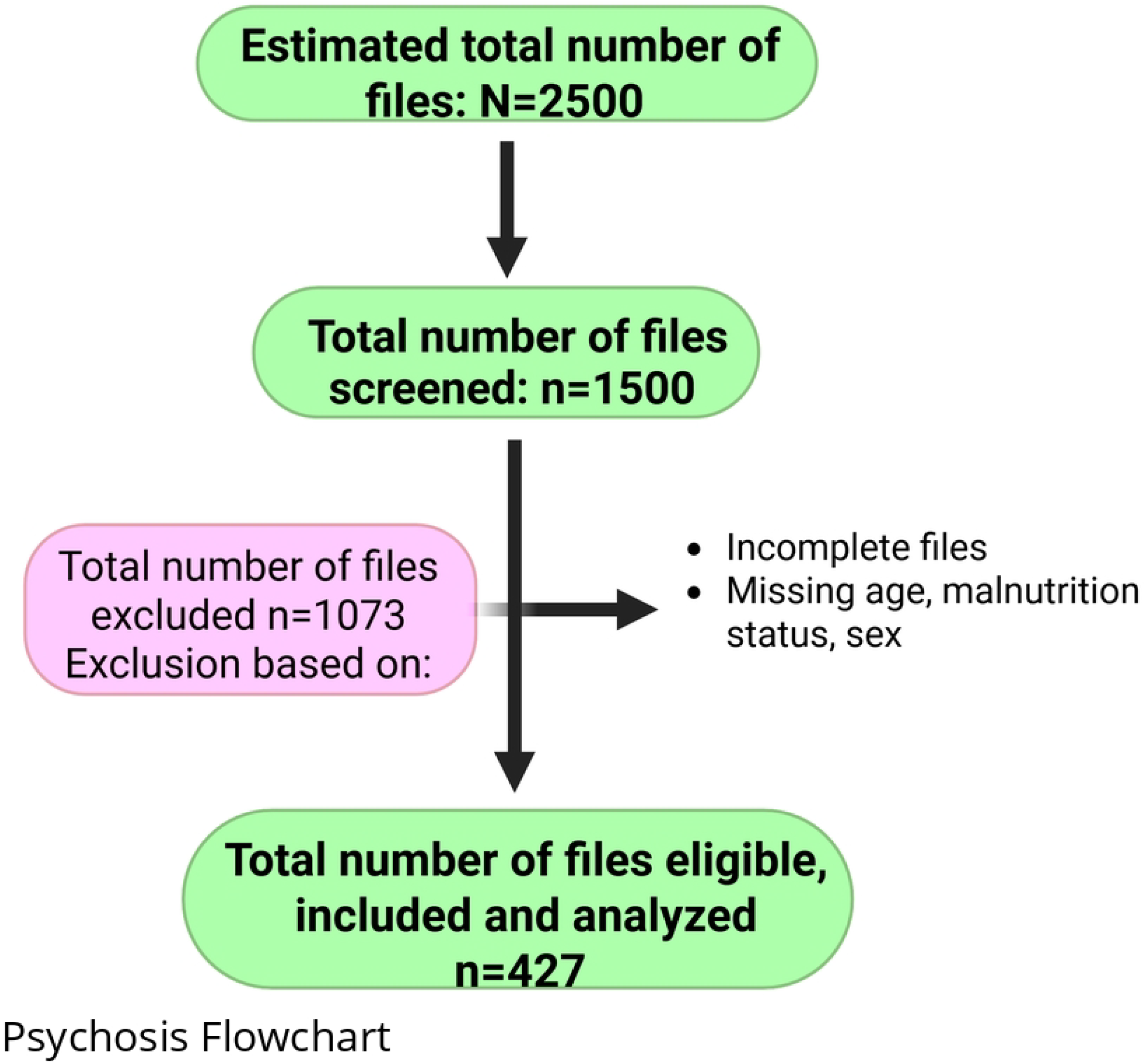
Eligibility flow diagram.

### Characteristics of the study population

This study enrolled a total of 427 participants. Male sex was strongly associated with the development of psychosis 84.0% (n=199). Urban residence was more prevalent among individuals with psychosis 51.8% (n=113). Meanwhile poor insight into mental illness was a prominent feature of psychosis 64.2% (n=129). With regards to substance use; majority of the participants were not on opioids 87.9% (n=210) while most of them were on cannabis, 56.5% (n=135). Additionally, majority of the participants did not have depression 90% (n=215) and anxiety 96.7% (231) (Table1).

**Table 1.**
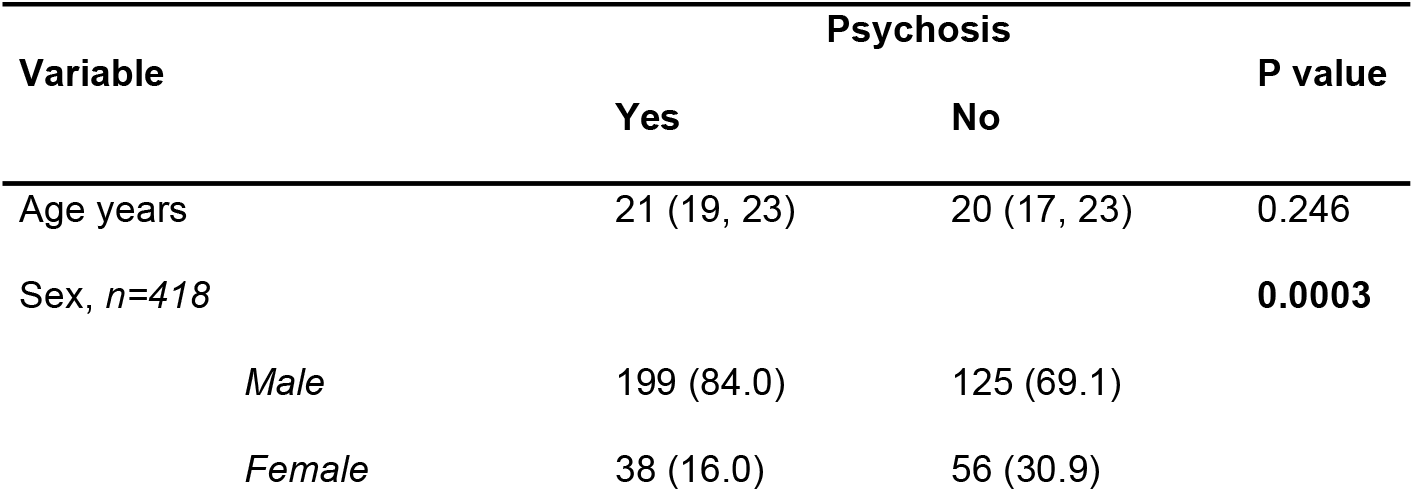
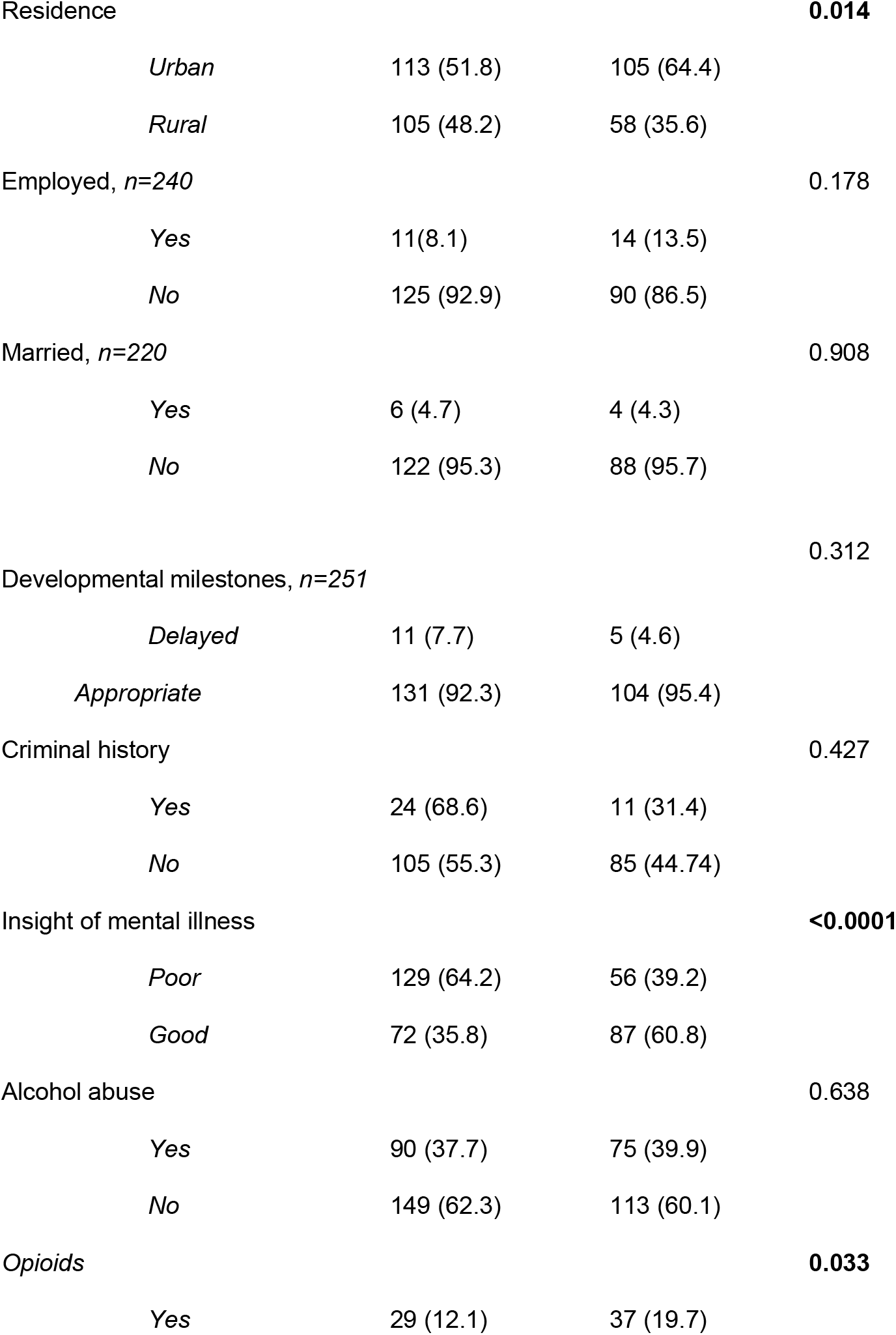
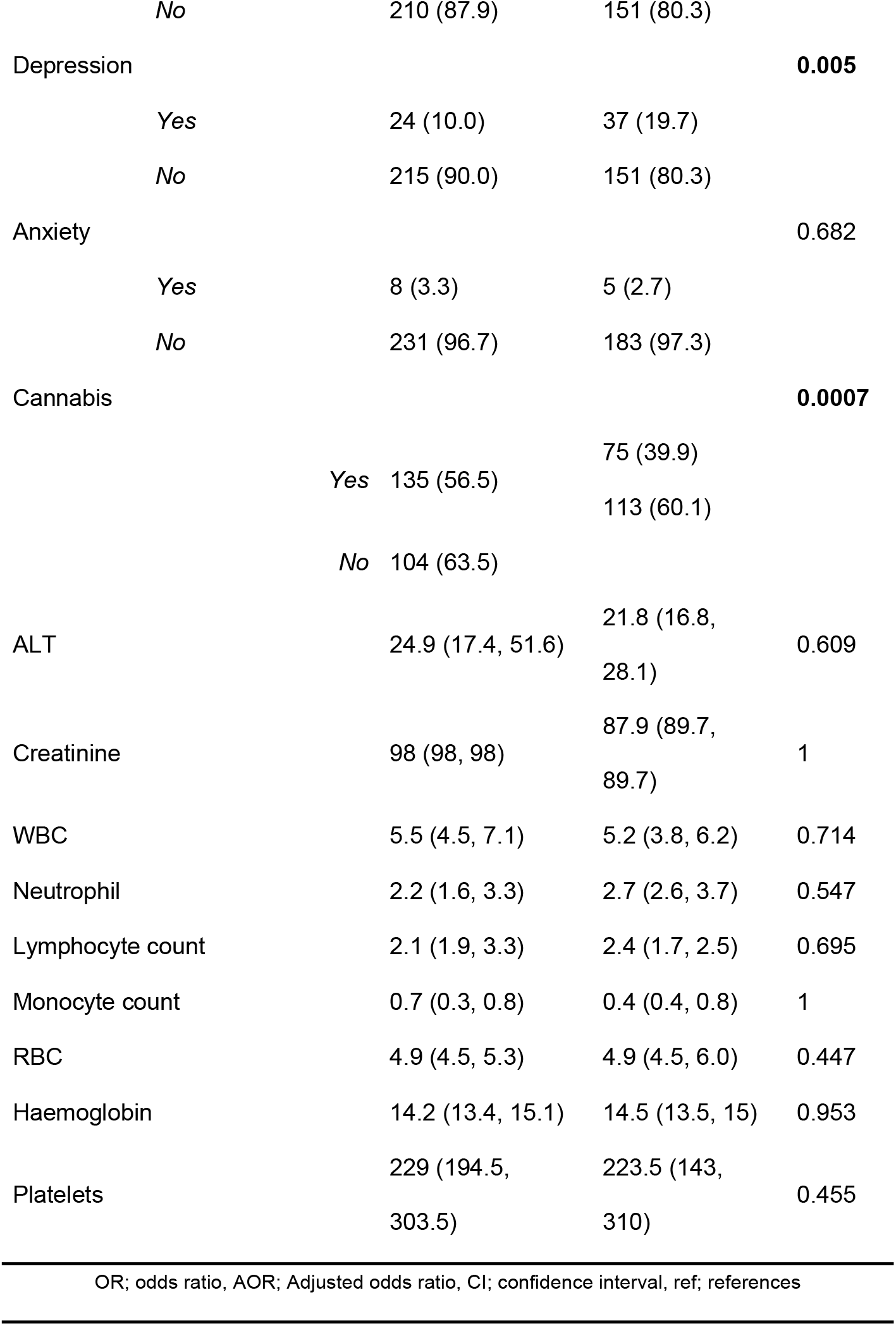
Characteristics of the study population.

| Variable | Psychosis |  | P value |
| --- | --- | --- | --- |
|  | Yes | No |  |
| Age years | 21 (19, 23) | 20 (17, 23) | 0.246 |
| Sex, n=418 |  |  | **0.0003** |
| *Male* | 199 (84.0) | 125 (69.1) |  |
| *Female* | 38 (16.0) | 56 (30.9) |  |
| Residence |  |  | **0.014** |
| *Urban* | 113 (51.8) | 105 (64.4) |  |
| *Rural* | 105 (48.2) | 58 (35.6) |  |
| Employed, *n*=240 |  |  | 0.178 |
| *Yes* | 11(8.1) | 14 (13.5) |  |
| *No* | 125 (92.9) | 90 (86.5) |  |
| Married, *n*=220 |  |  | 0.908 |
| *Yes* | 6 (4.7) | 4 (4.3) |  |
| *No* | 122 (95.3) | 88 (95.7) |  |
| Developmental milestones, *n*=251 |  |  | 0.312 |
| *Delayed* | 11 (7.7) | 5 (4.6) |  |
| *Appropriate* | 131 (92.3) | 104 (95.4) |  |
| Criminal history |  |  | 0.427 |
| *Yes* | 24 (68.6) | 11 (31.4) |  |
| *No* | 105 (55.3) | 85 (44.74) |  |
| Insight of mental illness |  |  | **<0.0001** |
| *Poor* | 129 (64.2) | 56 (39.2) |  |
| *Good* | 72 (35.8) | 87 (60.8) |  |
| Alcohol abuse |  |  | 0.638 |
| *Yes* | 90 (37.7) | 75 (39.9) |  |
| *No* | 149 (62.3) | 113 (60.1) |  |
| Opioids |  |  | **0.033** |
| *Yes* | 29 (12.1) | 37 (19.7) |  |

|  |  |  |  |  |
| --- | --- | --- | --- | --- |
|  | No | 210 (87.9) | 151 (80.3) |  |
| Depression |  |  |  | **0.005** |
|  | Yes | 24 (10.0) | 37 (19.7) |  |
|  | No | 215 (90.0) | 151 (80.3) |  |
| Anxiety |  |  |  | 0.682 |
|  | Yes | 8 (3.3) | 5 (2.7) |  |
|  | No | 231 (96.7) | 183 (97.3) |  |
| Cannabis |  |  |  | **0.0007** |
|  | Yes | 135 (56.5) | 75 (39.9) |  |
|  |  |  | 113 (60.1) |  |
|  | No | 104 (63.5) |  |  |
| ALT |  | 24.9 (17.4, 51.6) | 21.8 (16.8, 28.1) | 0.609 |
| Creatinine |  | 98 (98, 98) | 87.9 (89.7, 89.7) | 1 |
| WBC |  | 5.5 (4.5, 7.1) | 5.2 (3.8, 6.2) | 0.714 |
| Neutrophil |  | 2.2 (1.6, 3.3) | 2.7 (2.6, 3.7) | 0.547 |
| Lymphocyte count |  | 2.1 (1.9, 3.3) | 2.4 (1.7, 2.5) | 0.695 |
| Monocyte count |  | 0.7 (0.3, 0.8) | 0.4 (0.4, 0.8) | 1 |
| RBC |  | 4.9 (4.5, 5.3) | 4.9 (4.5, 6.0) | 0.447 |
| Haemoglobin |  | 14.2 (13.4, 15.1) | 14.5 (13.5, 15) | 0.953 |
| Platelets |  | 229 (194.5, 303.5) | 223.5 (143, 310) | 0.455 |

OR; odds ratio, AOR; Adjusted odds ratio, CI; confidence interval, ref; references

### Factors associated with Cannabis use in Logistic Regression

Based on the study participants, in univariable analysis; male individuals had lower odds of having psychosis compared to females (OR = 0.43 95% CI = 0.266–0.681, p = 0.0004) (Table 2). Urban residence was associated with higher odds of psychosis compared to rural residence (OR = 1.68, 95% CI = 1.109–2.551, p = 0.014). Poor insight to mental illness was strongly linked to reduced odds of psychosis (OR = 0.36, 95% CI = 0.230–0.559, p < 0.0001). Opioid use was associated with lower odds of psychosis (OR = 0.56, 95% CI = 0.331–0.956, p = 0.033). Depression was inversely associated with psychosis (OR = 0.46, 95% CI = 0.262–0.792, p = 0.0005). Cannabis use significantly increased odds of psychosis (OR = 1.96, 95% CI = 1.326–2.883, p = 0.0007).

**Table 2.**
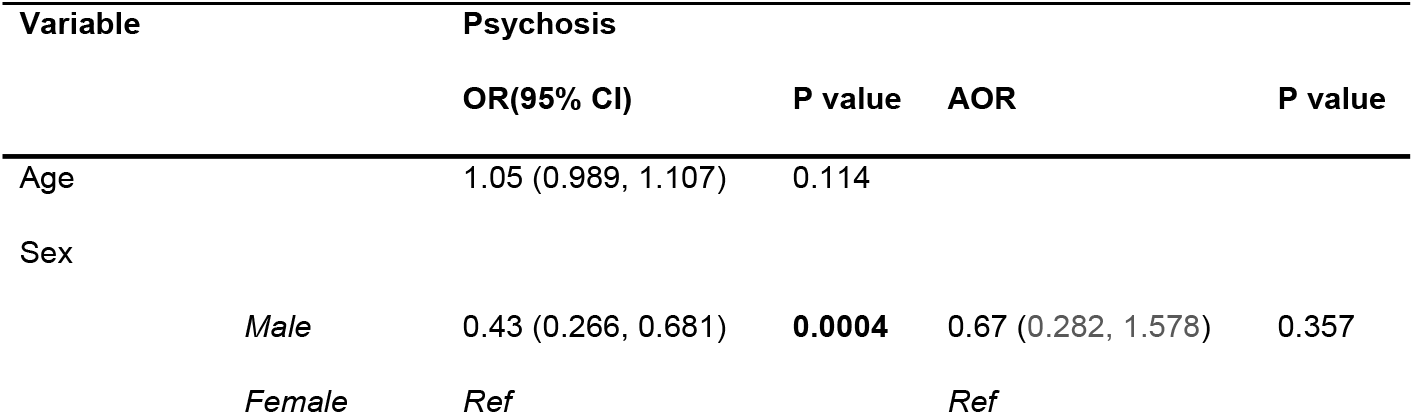
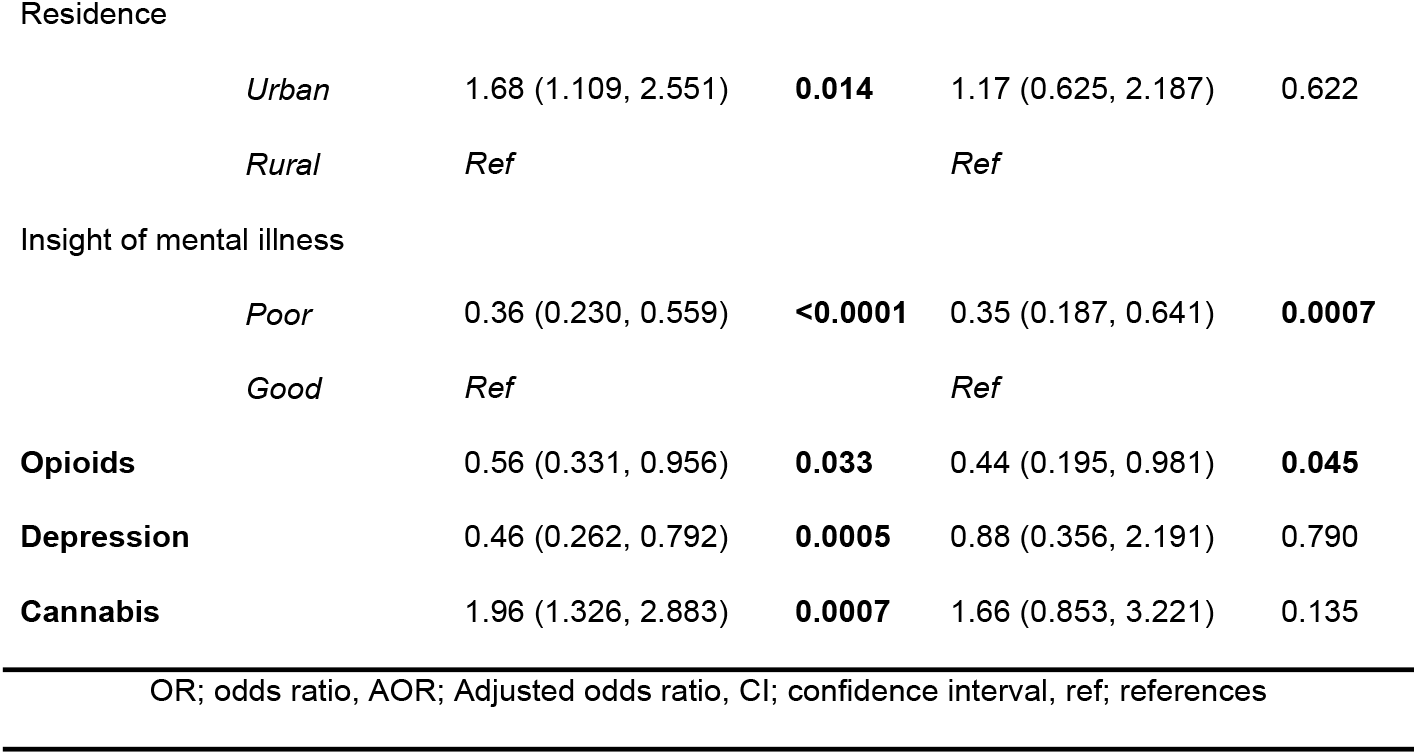
Factors associated with psychosis in Logistic Regression.

| Variable | Psychosis |  |  |  |
| --- | --- | --- | --- | --- |
|  | OR(95% CI) | P value | AOR | P value |
| Age | 1.05 (0.989, 1.107) | 0.114 |  |  |
| Sex |  |  |  |  |
| Male | 0.43 (0.266, 0.681) | **0.0004** | 0.67 (0.282, 1.578) | 0.357 |
| Female | Ref |  | Ref |  |

|  |  |  |  |  |  |
| --- | --- | --- | --- | --- | --- |
| Residence |  |  |  |  |  |
|  | *Urban* | 1.68 (1.109, 2.551) | **0.014** | 1.17 (0.625, 2.187) | 0.622 |
|  | *Rural* | *Ref* |  | *Ref* |  |
| Insight of mental illness |  |  |  |  |  |
|  | *Poor* | 0.36 (0.230, 0.559) | **<0.0001** | 0.35 (0.187, 0.641) | **0.0007** |
|  | *Good* | *Ref* |  | *Ref* |  |
| **Opioids** |  | 0.56 (0.331, 0.956) | **0.033** | 0.44 (0.195, 0.981) | **0.045** |
| **Depression** |  | 0.46 (0.262, 0.792) | **0.0005** | 0.88 (0.356, 2.191) | 0.790 |
| **Cannabis** |  | 1.96 (1.326, 2.883) | **0.0007** | 1.66 (0.853, 3.221) | 0.135 |
| OR; odds ratio, AOR; Adjusted odds ratio, CI; confidence interval, ref; references |  |  |  |  |  |

In multivariable analysis, only two factors retained significance: Poor insight to mental illness remained a predictor of lower odds of psychosis (AOR = 0.35, 95% CI = 0.187– 0.641, p = 0.0007). And opioid use continued to show reduced odds of psychosis (AOR = 0.44, 95% CI = 0.195–0.981, p = 0.045).

## Discussion

The study aimed to identify factors associated with psychosis in adolescents and young adults (aged 10–27 years) at LUTH. The study population was predominantly male (77.5%) and young with a median age of 21 years, reflects a high burden of psychosis among male adolescents and young adults in Zambia. While males constituted 84% of the psychosis group, the logistic regression paradoxically revealed that males had lower odds of psychosis compared to females. This discrepancy may stem from the overwhelming male predominance in the sample size (77.5%), which could skew representation. Similar male-dominated cohorts have been reported in African psychiatric studies, potentially due to cultural factors influencing help-seeking behaviour, where males with severe symptoms (e.g., aggression) are more likely to be hospitalized [2,3]. Our findings are in agreement with [29] which found that males comprised 72% of psychosis cases. However, the inverse association between male sex and psychosis in regression models contrasts with Western studies, where male sex is a well-established risk factor for schizophrenia [30]. This divergence may reflect differences in diagnostic practices, cultural perceptions of mental illness, or distinct etiological pathways in low-resource settings like Zambia[12].

Poor insight into mental illness emerged as the strongest independent predictor of psychosis, consistent with [19,31,32] where poor insight was prevalent in 60–70% of psychosis cases. Poor insight often delays treatment-seeking and exacerbates stigma, particularly in settings with limited mental health literacy. Our findings reinforce the need for community-based psychoeducation programs to improve early recognition of psychotic symptoms. However, the inverse association (poor insight linked to reduced psychosis odds) is counterintuitive and may reflect methodological nuances, such as insight being assessed post-diagnosis, potentially conflating cause and effect.

Opioid use was associated with reduced psychosis. This contrasts sharply with global literature, where opioids are rarely protective [5,33]. One plausible explanation is self-medication: individuals with psychotic symptoms may avoid opioids due to heightened paranoia or sedation. Alternatively, opioids’ neurobiological effects (e.g., dopamine modulation) might transiently ameliorate psychotic symptoms, as hypothesized in [34,35]. However, this finding warrants caution, as opioid use in Zambia is often illicit and linked to comorbid risks (e.g., HIV, overdose) that were not assessed here.

### Study limitations and strengths

#### Strengths

Our study offers valuable insights into psychosis among Zambian youth, with several strengths. Relatively we had a large sample size (n=427) enhances statistical power, and the inclusion of both sociodemographic and clinical variables provides a comprehensive analysis. The use of multivariable regression adjusts for confounders, strengthening causal inferences for factors like poor insight and opioid use. Additionally, focusing on an understudied population in a low-resource setting addresses a critical gap in global mental health research. The robust association between cannabis use and psychosis in univariable analysis aligns with global evidence, underscoring substance use as a modifiable risk factor.

#### Limitations

The cross-sectional design precludes causal conclusions, and the single-center sample limits generalizability. Self-reported substance use risks recall bias, potentially underestimating true prevalence. The male-dominated cohort (77.5%) may skew findings, particularly as male sex paradoxically showed reduced psychosis odds in regression. Furthermore, key variables like urban residence and cannabis use lost significance in adjusted models, suggesting unmeasured confounding (e.g., socioeconomic factors). Finally, the lack of validated tools for assessing insight or psychosis severity may affect measurement accuracy. Despite these limitations, the study provides foundational data for future longitudinal and intervention-focused research in similar settings.

## Conclusion

This study highlights critical sociodemographic and clinical factors linked to psychosis among Zambian youths. Male sex, rural residence, poor insight, reduced opioid use, and higher cannabis use were key correlates. While cannabis use showed a strong univariable association with psychosis, its effect attenuated after adjustment, suggesting confounding by insight or substance interactions. Conversely, poor insight and opioid use emerged as independent predictors, underscoring their clinical relevance. The paradoxical protective role of opioids warrants further exploration. These findings emphasize the need for context-specific interventions, including mental health literacy programs and substance use harm reduction strategies. Future research should employ longitudinal designs to clarify causal pathways and address limitations like cross-sectional bias. By bridging gaps in African mental health data, this study informs targeted policies to mitigate psychosis burden in low-resource settings.

## Data Availability

The raw data underlying the results presented in the study have been uploaded as supporting information.

## Supplementary files

S1. Strobe checklist

S2. Data

